# Data Imputation for Clinical Trial Emulation: A Case Study on Impact of Intracranial Pressure Monitoring for Traumatic Brain Injury

**DOI:** 10.1101/2023.01.29.23285172

**Authors:** Zhizhen Zhao, Ruoqi Liu, Jonathan I. Groner, Henry Xiang, Ping Zhang

## Abstract

Randomized clinical trial emulation using real-world data is significant for treatment effect evaluation. Missing values are common in the observational data. Handling missing data improperly would cause biased estimations and invalid conclusions. However, discussions on how to address this issue in causal analysis using observational data are still limited. Multiple imputation by chained equations (MICE) is a popular approach to fill in missing data. In this study, we combined multiple imputation with propensity score weighted model to estimate the average treatment effect (ATE). We compared various multiple imputation (MI) strategies and a complete data analysis on two benchmark datasets. The experiments showed that data imputations had better performances than completely ignoring the missing data, and using different imputation models for different covariates gave a high precision of estimation. Furthermore, we applied the optimal strategy on a medical records data to evaluate the impact of ICP monitoring on inpatient mortality of traumatic brain injury (TBI). The experiment details and code are available at https://github.com/Zhizhen-Zhao/IPTW-TBI.

## Introduction

Clinical trials are essential for evaluating the safety and efficacy of treatments for diseases. Clinical trials could inform researchers how the treatment will interact with human that cannot be learned in the laboratory or in animals. However, clinical trials, especially randomized clinical trials (RCTs) are often expensive in financial costs and execution time. Considering the size and limitation of clinical trials, sometimes it’s difficult to generalize results to the larger and more inclusive populations in practice. Real-world data (RWD) refers to observational data in health care derived from the conventional clinical environment, such as electronic health records (EHRs), medical claims, billing activities and product records^1^. Understanding the real-world evidence from RWD can help complement the knowledge obtained from the clinical trials. In addition, emulating RCTs using RWD is very popular, which could help assess the safety and effectiveness of a treatment before running a large-scale RCT and provide guidance on the design of more-inclusive trials. For example, Stewart et al. extracted the real-world intermediate end points data from different health care data organization and found the consistency between these observed data with similar end points and outcomes in clinical trials for immunotherapy-treated advanced non-small-cell lung cancer^2^. Chen et al. emulated a RCT of Alzheimer’s disease using RWD, generated the results comparable to the original RCT, and explored the potential problem of generalizability of the RCT^3^.

However, missing data are challenges in use of real-world observational data, which could cause substantial loss of information^4^. If missing data are not handled properly, it would lead to bias when estimate causal effect of treatments. Missingness could be caused by several mechanisms^5^. In this study, we focused on missing at random (MAR), that is conditional on other observed information, the probability of missing data for a subject is independent of the values themselves^6^. Previous literature proposed three strategies to handle missing data^4^. The easiest method is to conduct a complete case analysis, that is to ignore all subjects with missing value. This is favorable when only a small number of data is missing^7^, otherwise it would waste costly collected data and lead to a biased and invalid conclusion. The second method is to drop variables with missing values, that is to only analyze the variables with complete data. However, it is infeasible if the interested variable has some missing data. The other approach is called multiple imputation (MI), which uses all observed data to fill in plausible values for the missing values^4–6,8,9^. Multiple imputed data sets are generated to consider the uncertainty in the imputation. Further analysis is performed on each imputed data set separately and estimators and variances are calculated by combining the results from multiple imputed data sets^5^. Compared with MI, the first two strategies might lead to a biased estimation, decreased coverage rate of confidence interval and the decreased discriminative ability of the multivariable model^10^. However, few literature investigated these strategies in the estimation of medical treatment effect. Berrevoets et al. found that selective imputation performed better than naively impute all data in causal context^11^, but it’s difficult to identify either missingness causing treatment or missingness caused by treatment pattern in practice. Thus our study aimed to compare various MI strategies with complete case analysis in treatment effect estimation systematically to evaluate whether imputing data works better and which imputation strategy should be adopted in RCT emulation using RWD.

The objective of this study was to select one proper methodology to deal with the missing data issue in the estimation of treatment effect in the observational study based on medical records. Since the ground truth of treatment effect in RWD is not available, we performed four different imputation strategies and a complete data analysis on two benchmark datasets to estimate the average treatment effect (ATE) through inverse probability of treatment weighting (IPTW). We found that imputing data to fill in missing values gave a higher precision in ATE estimation than deleting all observations with missing values. The choices of imputation models did not show significant differences, but overall using a combination of different models for different covariates to create imputed data performed the best. In addition, we conducted a case study to evaluate the effect of intracranial pressure (ICP) monitoring on in-hospital mortality among patients with severe traumatic brain injury (TBI). Record-level National Trauma Data Bank (NTDB) data for patients with TBI from 2017 to 2018 were analyzed in the study^12^. We applied the selected imputation method to replace the missing data in the original observational dataset. The distribution of imputed data was similar to the complete data but kept all patients’ records. On the basis of imputed data, we performed IPTW model to adjust the baseline characteristics of patients and then inferred causal relations in terms of ATE. All covariates were balanced well after weighting with all absolute standardized differences (ASDs) less than the threshold, 0.1^13^. Finally we estimated the ATE of ICP monitoring and inferred the causal conclusion that treated by ICP would lower the in-hospital mortality rate among severe TBI patients.

Overall, our contributions can be summarized as follows: (1) We compared various strategies to handle the missing data problem in causal analysis, either removing observations with missing values or generating multiple imputed data to fill in missing values. MI can help make full use of the real-world data. We combined these methods and IPTW model to adjust confounders and estimate ATE. (2) We performed the proposed methodologies on two benchmark datasets. The results indicated that the estimation performances based on the imputed data were much better than conducting a complete data analysis. Missing data imputation is a proper way to handle missingness in the estimation of ATE when missing rates are large. (3) We applied the MI strategy (combination of models) on the observational NTDB dataset and analyzed the causal effect of ICP monitoring on mortality. Our study is the first to conduct the causal analysis using a large-scale data of severe TBI patients, and the results complemented the knowledge in this field. We applied the IPTW to remove the confounding effects. The high degree of balance was achieved in the pseudo patient population so that we could infer a valid causal conclusion. Finally, we found ICP monitoring is helpful in reducing the mortality significantly among severe TBI patients.

### Methodology

**Setup** Let *X* = (*X*_1_, *X*_2_, …, *X*_*K*_) denotes a vector of complete *K* covariates. *X* is a partially observed random sample from the multivariate distribution *P* (*X*|*θ*), where we assume the multivariate distribution of *X* is completely specified by a vector of unknown parameters, *θ. p* = (*p*_1_, *p*_2_,…, *p*_*K*_) denotes a vector of missing rate for each covariate. *Z* represents the treatment where denote *Z* = 1 as being treated (in the treated group) and *Z* = 0 as untreated (in the control group). *Y* represents the observed outcome. In this study, we examined the scenario of binary treatment and binary outcome, but the methodology can also be extended to multi-level treatment and continuous outcomes. Our goal is to select an optimal strategy to deal with missing data when estimate ATE.

### Multiple Imputation for Missing Data

Multiple imputation is valid under the assumption of MAR that missing data are related to other available characteristics and are random conditional on other observed variables. MI imputes missing values multiple times through random draws from distributions inferred from available data. We adopted one popular approach, multiple imputation by chained equations (‘MICE’)^14,15^, to fill in missing data in baseline covariates. The analysis of imputed data consists of three steps shown in Figure1: generate *m* imputed datasets, analyze the imputed data, and pool the analysis results^16^.

**Figure 1:**
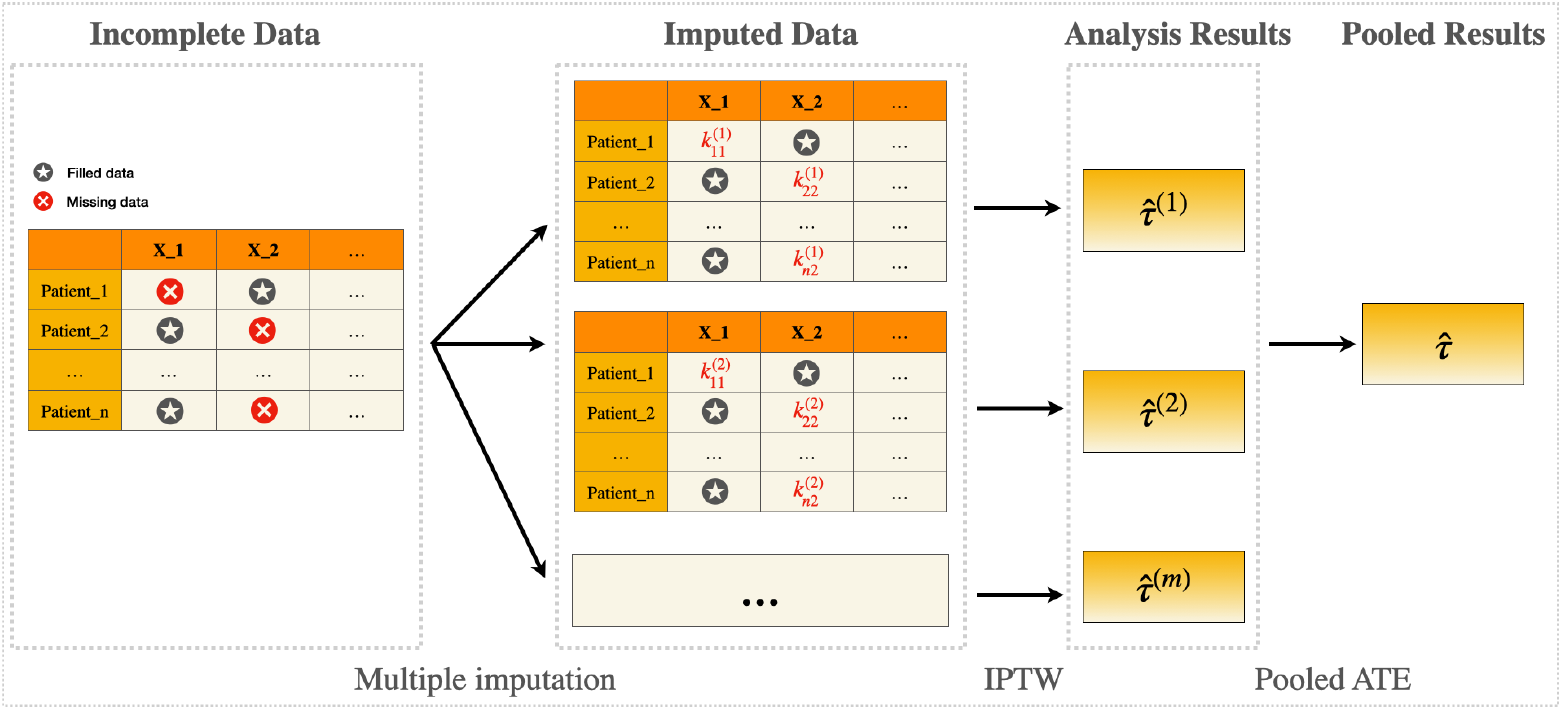
Three steps used in the analysis of imputed data: The first step is to create multiple *m* imputed datasets where missing values are replaced by *m* plausible values through random draws. 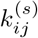 represents the imputed data for the (*i, j*) missing value during the *s*^*th*^ iteration, where *s* = 1, …, *m*. Next is to estimate ATE 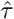 for each imputed dataset by IPTW separately. The last step is to pool the results obtained in each imputed dataset together to arrive at a single ATE estimation 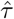.

For the generation of multiple imputation, this approach specifies univariate predictive distribution of each variable of missingness given all other observed variables and then repeatedly sample data from the conditional distribution. Each incomplete covariate has its own imputation model. For the *i*^*th*^ covariate *θ*_*i*_, the MICE algorithm obtained its posterior distribution by sampling iteratively from the conditional distribution *P* (*X*_*i*_ |*X*_*−i*_, *θ*_*i*_), where *X*_*−i*_ denotes all other covariates except *X*_*i*_. In the first iteration, it is draw from the observed marginal distributions. Then, for the *j*^*th*^ iteration, it is a Gibbs sampler that 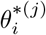draws from 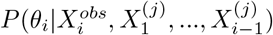 and 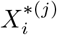 draws from 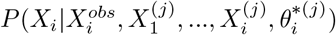, where 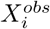 is the observed value in the *i*^*th*^ covariate and 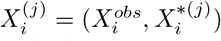 is the *i*^*th*^ imputed value at the *j*^*th*^ iteration. The process is repeated for all incomplete variables multiple times to impute one complete dataset. The whole process is repeated *m* (*m* = 5) times to generate *m* imputed datasets. Then perform IPTW on each imputed dataset to obtain weights, estimate ATE and pool the results.

In order to find one proper way to impute data in RWD, we compared different imputation models in MICE:(1) predictive mean matching (pmm) and logistic regression (logreg) model to impute values for continuous and categorical variables; (2) random forest (rf) model to impute values; (3) classification and regression trees (cart) model to impute values; (4) combination of predictive mean matching, logistic regression, random forest and classification and regression trees model to impute values, where pmm for continuous variables, all other methods randomly assigned to the same number of categorical variables.

Predictive mean matching is the most conventional way for MICE by fitting a linear regression model for the observed data and predict the conditional mean for the missing data. But this method faces the problem brought by the nonlinear term, interaction, and collinearity^17^. Classification and regression trees and random forest do not depend on the distribution assumptions and can better accommodate these issues. Besides MI, we also conducted a complete data analysis where all subjects with missing values were removed and IPTW was performed on the left available subjects.

### Treatment effect Estimation

#### Assumptions

The proposed method is used to evaluate the causal effect of using ICP monitoring on in-hospital mortality for severe TBI patients. Causal inference analysis relies on the potential outcome framework and the definition of potential outcomes relies on the stable unit treatment value assumption (SUTVA)^18^. Under the binary treatment assignment, every subject (the *i*^*th*^ unit) has a pair of the potential outcomes 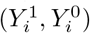. However, in reality, only one of two potential outcomes could be observed. The observed outcomes can be connected to the potential outcomes through the equation 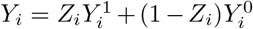. For binary outcome, ATE can be viewed as the difference between random variable *Y* ^1^ and *Y* ^0^. It is also known as the risk difference: *τ* = *E*[*Y* ^1^] −*E*[*Y* ^0^] = *P* (*Y* ^1^ = 1) −*P* (*Y* ^0^ = 1), where *E*[*Y* ^*z*^] denotes the potential expected prevalence of patients who would have died during the follow-up period if all patients in the study had been assigned with treatment *z*. In the observational study, the treatment assignment mechanism is unknown and usually far from random. Confounders (pre-treatment covariates) would affect both treatment assignment and outcomes. Thus we need to assign weights to patients in two groups to estimate the population quantities to adjust confounders.

### Inverse Probability of Treatment Weighting

Covariate balance has been shown to be important to estimate unbiased causal effects^19,20^. Propensity score based methods are the most popular way to achieve covariate balance^21^. The propensity score, *e*(*x*), is defined as the conditional probability of exposure to treatment, i.e monitored by ICP, given the observed covariates^22^. The propensity score is usually estimated by running a binomial logistic regression model where the outcome variable is 1 if the patient received the treatment and 0 if the patient did not receive the treatment. All baseline characteristics would be included in the propensity score model.

We performed inverse probability of treatment weighting to adjust confounders. This method is to weight each subject based on propensity score to create a pseudo dataset where the baseline covariates are independent of the treatment assignment^21^. Then the causal effect was analyzed with conventional regression model. For each patient, the weight is equal to the inverse of the probability of receiving the treatment that the patient actually received. Weights are defined as 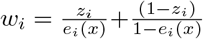 for the subject *i*. However, the weights can become very large for treated subjects with very low propensity scores or untreated patients with very high propensity scores. If some subjects had extremely large weights, the resulting ATE estimator would have a large variance and was not approximately normally distributed. We addressed this issue by using stabilized weights instead, where stabilized weights were defined as 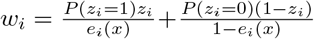 for the subject *i*, and the numerator is the marginal probability of receiving the actual treatment received instead of 1.

### Estimation of ATE

For simplicity, we used the treatment-only regression model to estimate ATE. For the imputed data, *m* imputed samples were created and the estimators were the pooled results in these samples. In the case study, we hypothesized the ICP was effective in lowering the mortality rate in TBI patients if the estimated ATE is negative, that is mortality rate in the treated group was significantly lower than the value in the control group at the confidence level of 95%.

### Experiment and Results Benchmark Datasets

**Twins** The semi-synthetic twins dataset was obtained from all twins birth in the USA between 1989 and 1991^23^. We created a binary treatment variable *Z* so that *Z* = 1 denotes being born the heavier and *Z* = 0 denotes being born the lighter. The outcome was identified as the mortality of each of the twins after one year. After eliminating records with missing values, the dataset contained 64,240 subjects with 40 pre-treatment covariates. Since information for both twins were available, outcomes within a pair could be considered as two potential outcomes and thus the true individual treatment effect was available. We adopted the following procedure to selectively choose one of twins as the observation and hide the other to create selection bias: *Z*_*i*_|*x*_*i*_ ∼ *Bernoulli*(*Sigmoid*(*w*^*t*^*x* + *ϵ*)), where *w*^*t*^ ∼ *Uniform*((−0.1, 0.1)^40*∗*1^) and *ϵ* ∼ *Noraml*(0, 0.1)^24^.

**IHDP** Hill compiled the dataset based on the Infant and Development Program(IHDP)^25^, where the covariates were obtained from a randomized experiment studying the effects of specialist home visits on cognitive test scores. The dataset contained 747 subjects with 25 covariates, where 139 as treated and 608 as control. We used the emulated outcome implemented as setting ‘A’ in the NPCI package^26^. In order to create a dataset similar to the data in our case study, we transformed the outcome into binary outcome by coding the top 35.9% of values as 1 and others as 0.

### Experiment Setup

The counterfactual outcomes and the ground truth effect are often missing in practice, so it is difficult to evaluate the ATE estimation directly on the observational datasets. In order to evaluate various MI strategies, we conducted two experiments on two semi-synthetic datasets, Twins and IHDP, with different settings. In the two datasets, we randomly selected half of covariates and created missing values under MAR mechanism. In the first experiment, the missing rate for each covariate was uniformly chosen between 0 and 80%. In the second experiment, the missing rates of selected covariates were consistent with the rates in our case study NTDB dataset, where detailed information can be found below. In the simulated datasets, 30% to 40% values were missing for 25% of covariates, 20% to 30% values were missing for 15% of covariates, 1% to 10% values were missing for 40% of covariates, and less than 1% values were missing for 20% of covariates. Figure 2 shows the missing rates of missing variables in one simulated data. In both experiments, the complete data analyses were performed by deleting all missing observations and various MI strategies were performed by using four different imputation models. All processes were repeated 1,000 times.

**Figure 2:**
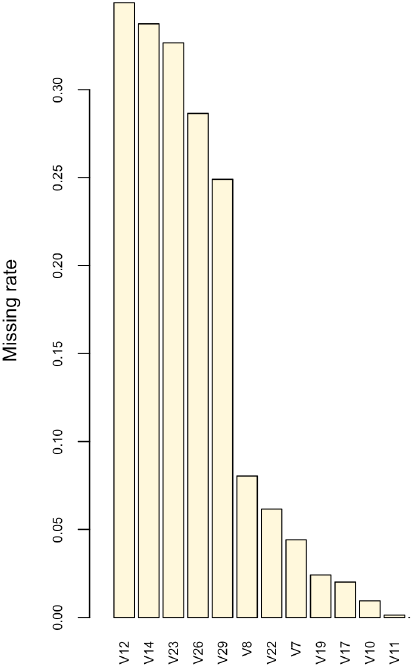
Missing rates in a simulated data in Experiment 2. The x-axis is missing variable and y-axis is missing rate of each variable.

**Performance metrics** We compared these strategies with respect to ATE estimation. For two benchmark datasets with ground truth, we used the expected precision in the estimation of ATE as a performance metric in evaluations: 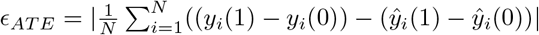, Where 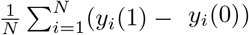 is the ground truth of the treatment effect and 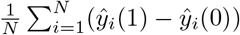 is the estimated treatment effect. The smaller value represents the better the performance. For the observational data, covariate balance across the treated and control groups in the pseudo-population generated by IPTW was assessed using the absolute standardized differences (ASDs)^22^. For continuous variables, it is defined as

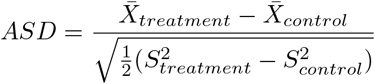

where 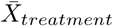 and 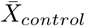 denote the sample mean of the covariate in two groups;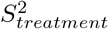 and 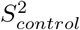 denote the sample variance of the covariate in two groups. For dichotomous variables, it is defined as

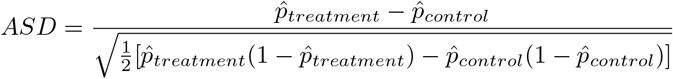

where 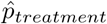 and 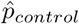 denote the prevalence of the dichotomous variable in two groups. Values below 0.1 are considered negligible differences^27^.

### Results on Benchmark Datasets

The performance of two experiments is reported in Table 1. The complete case analyses show the worst performance on both datasets with the highest value of *ϵ*_*AT E*_ and standard deviation. All MI strategies achieved much better performances. These results confirmed that imputing the missing data could estimate the counterfactual outcome and ATE better. Compared with experiment 1, the gap between complete case analysis with other MI strategies in experiment 2 was reduced. This is might because the overall missing ratio in experiment was smaller, with the highest missing proportion in one covariate being 40%, meaning that fewer cases were deleted in experiment 2. This also confirmed our expectation that fewer missing values would make the estimation more precise. Various multiple imputation strategies demonstrated similar performances. One possible reason is that the ATE estimation highly relies on the balance of pre-treatment confounders. When we performed IPTW model on each imputed dataset, propensity score was estimated to assign weight to each patient. In this process, the issues such as interaction and collinearity relationship between covariates might not affect too much if the covariates were well balanced. The combination of different strategies for different type of variables had a slightly higher precision in both datasets. Thus we would perform this MI method in the following case study to analyze the effect of ICP monitoring on in-hospital mortality.

**Table 1:** Performance comparisons on Twins and IHDP. The missing rates were randomly selected in Experiment 1. The missing rates were consistent with the rates in our case study in Experiment 2.

| Method | Experiment 1 |  |  |  | Experiment 2 |  |  |  |
| --- | --- | --- | --- | --- | --- | --- | --- | --- |
|  | Twins |  | IHDP |  | Twins |  | IHDP |  |
| | $\epsilon_{ATE}(\%)$ | SD | $\epsilon_{ATE}$ | SD | $\epsilon_{ATE}(\%)$ | SD | $\epsilon_{ATE}$ | SD |
| Complete case analysis | 0.5963 | 0.01577 | 0.0804 | 0.20067 | 0.1592 | 0.00206 | 0.0151 | 0.03791 |
| MI: pmm+logreg | 0.0512 | 0.00025 | **0.0132** | 0.00203 | 0.0445 | 0.00018 | 0.0134 | 0.00130 |
| MI: rf | 0.0514 | 0.00024 | 0.0134 | 0.00209 | 0.0442 | 0.00016 | 0.0135 | 0.00138 |
| MI: cart | **0.0486** | 0.00023 | 0.0135 | 0.00207 | 0.0425 | 0.00017 | 0.0135 | 0.00132 |
| MI: combination | 0.0488 | 0.00024 | **0.0132** | 0.00200 | **0.0420** | 0.00018 | **0.0133** | 0.00116 |

### Case Study: Effect of ICP Monitoring on Traumatic Brain Injury Inpatient Mortality

**Background** Traumatic brain injury is the leading cause of death and disability^28^. In the US, around 1.5 million people sustain TBI. More than 50,000 Americans died and approximately 500,000 suffered permanent neurological sequelae as a result of TBI^29^. ICP monitoring for patients with severe TBI has been recommended in the Brain Trauma Foundation(BTF) published management guidelines since the 1970s. However, few RCTs have been conducted successfully to examine the effect of ICP monitoring on severe TBI patients’ outcomes. The difficulties include the ethical concern of making a control group where patients are not monitored by ICP, and the widely use of ICP monitoring as a treatment in major hospitals. Although a few researchers studied the associations of ICP-based treatment with patients’ recovery^28,29^, little research focuses on the causal relationship between ICP monitoring and outcomes. In this case study, our goal was to evaluate the causal effect of ICP monitoring on in-hospital mortality.

### Study Design

Total Glasgow Coma Scale (GCS) score is often used to define the severity of TBI and a GCS score of 8 or less defines a severe injury^30^. In order to investigate the effect of ICP monitoring among severe TBI patients, the patients in the study were selected to satisfy the inclusion criterion: Total GCS score ≤8. The treatment considered was the use of ICP monitoring. Cerebral monitors include intraventricular drain/catheter, intraparenchymal pressure monitor, intraparenchymal oxygen monitor and jugular venous bulb. Patients were considered as treated if at least one monitoring was used during treatment (coded as 1) and untreated if no monitoring was used (coded as 0). The index time was defined as the time of starting ICP monitoring for treated patients. The baseline period was defined as the time period prior to the index time, that was from obtaining the information for patients in the scene to the time of deciding if patients receiving the ICP monitoring or not. Baseline data were used to characterize the patients before taking the treatment. The follow-up period started from the index time until discharge from hospital. The outcome was defined as mortality and measured during the follow-up period. The patient was considered to have the outcome if he or she died in hospital. Confounders were defined as variables that could affect treatment assignments and outcome. We consulted TBI domain experts to compile a list of 18 hypothesized confounding factors (shown below) which might affect the treatment assignment and outcome and were measured during the baseline period.

### Observational NTDB Dataset

The Trauma Quality Programs (TQP) Participant Use File (PUF) is a set of relational tables containing elements as defined by NTDB for each respective admission year. It is an incident-based database from a voluntary subset of trauma centers, which provides valuable real-world patient data for trauma care quality assessment and research. However, a substantial proportion of patients (36%) had missing values in one or more risk factors. Analyzing the subjects with available data only would waste one third of precious data. Therefore we decided to handle the missingness problem and built the imputation model. The dataset included 572,185 traumatic records (admitted between 2017 and 2018) with more than 70 variables of patients in demographic information, pre-hospital data, emergence department data, measures for the process of cares. To evaluate the effect of ICP monitoring on mortality among patients with severe traumatic brain injury, the interested outcome in the analysis was hospital mortality. The disposition ‘deceased/expired’ would be coded as died (1) and others as survived (0) when discharged from the hospital. Pre-treatment confounders include age, transportation mode, initial EMS systolic blood pressure, initial EMS pulse rate, EMS total GCS, etc, where 6 were categorical variables and 12 were continuous variables. The dataset was available by request^12^. All detailed variables and their missing rates could be found in https://github.com/Zhizhen-Zhao/IPTW-TBI.

Considering the inclusion criteria, only 74,819 patients with total GCS score less than 9 were included in the analysis. Among them, 5,261 (7.03%) and 10,879 (14.54%) patients did not have information about treatment and outcome, respectively. After we removed these patients, 60,640 patients remained in the dataset, in which 13,951 (23.01%) were in the treated group and 46,689 (76.99%) were in the control group. The raw in-hospital mortality rates without adjustment were 53.97% and 56.34% for treated and control patients, respectively. We only considered pre-treatment covariates and remove other variables (measured after index time) in the data set. Among baseline covariates, 15 variables had missing values where one third of them had more than 10% of missing values, with the highest missing rate being 37%.

### Results on NTDB Dataset

For our interested patients, 21,810 (36.97%) had at least one missing value, suggesting that if we did not perform MI, nearly 40% of data would be wasted. On the basis of previous experiments, we used the common method MICE to impute missing values where using pmm for continuous variables, rf, cart and logreg for categorical variables. Figure 3 reports the distributions of observed data and imputed data for continuous variables. The matching shapes showed that the distributions were very closed and the imputed values were plausible. Thus we could use the imputed data set for further analysis.

**Figure 3:**
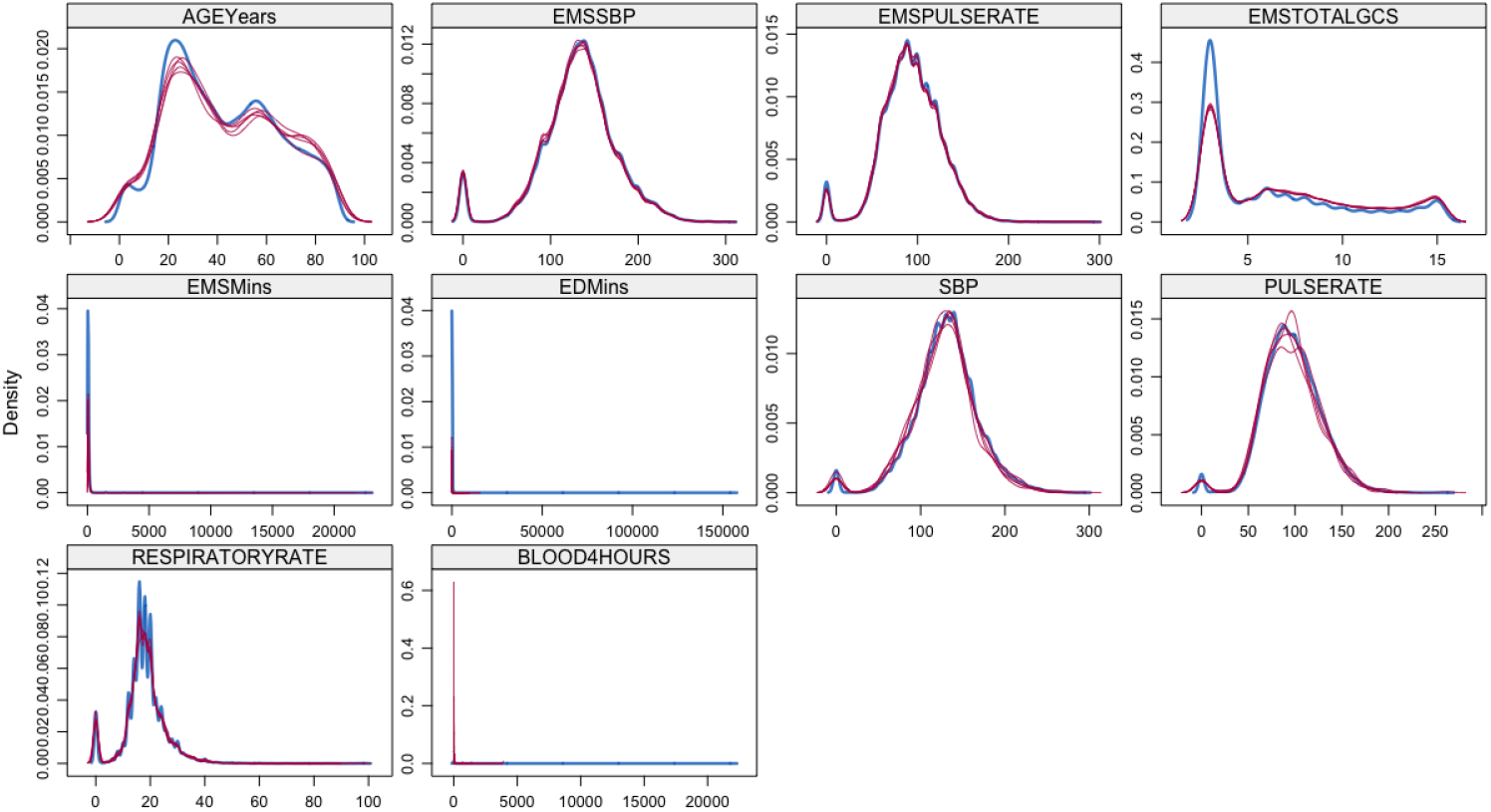
Density plots of imputed variables: the density of imputed data for each imputed data set is showed in megenta; the density of observed data is showed in blue.

### Covariate Balance

We applied the proposed IPTW model to the data. The objective was to generate a balanced sample to properly assess causal differences in mortality across two groups. We assigned weights to each patients on the basis of propensity score, which was estimated using binomial logistic regression. We examined if covariates were fully balanced between two groups after weighting through ASD. Table 2 reports the sample mean for continuous covariates in treated and control groups before weighting as well as the ASD values before and after weighting. Figure 4 presents the visualization of ASD value of each covariate before and after weighting. The original unadjusted data were denoted as red dots and the IPTW weighted pseudo data were denoted as blue triangles. One third of variables in the original dataset had ASD values above 0.1^13^, indicating that the original observational data were biased and potential confounding variables existed. The largest ASD value is 0.284 in the AGEYears. However, all ASDs were below 0.1 in the imputed dataset by IPTW, with the largest single value being 0.014. These results indicated good balance across all covariates. Thus, the estimation of ATE based on the pseudo-patient-population would be more valid and accurate than the original data.

**Table 2:** ASDs for weighted and unweighted samples in case study

| Variable | Control Group* | Treated Group* | ASD (unweighted) | ASD (weighted) |
| --- | --- | --- | --- | --- |
| Age | 44.285 | 38.056 | 0.284 | 0.006 |
| EMSSBP | 132.187 | 132.464 | 0.006 | 0.003 |
| EMSpulserate | 93.510 | 94.201 | 0.022 | 0.001 |
| EMStotalGCS | 6.597 | 5.706 | 0.227 | 0.006 |
| EMSMins | 79.404 | 69.716 | 0.037 | 0.005 |
| EDMins | 104.513 | 101.413 | 0.004 | 0.004 |
| SBP | 130.641 | 131.761 | 0.030 | 0.001 |
| Pulserate | 97.189 | 98.297 | 0.037 | 0.005 |
| Respiratoryrate | 18.172 | 18.778 | 0.080 | 0.001 |
| TotalGCS | 4.160 | 4.063 | 0.056 | 0.014 |
| Blood4hours | 65.491 | 83.347 | 0.034 | 0.004 |
| Bedsizes | 3.028 | 3.014 | 0.015 | 0.001 |

\*Numbers in the second and third columns are sample means for continuous variables before weighting.

**Figure 4:**
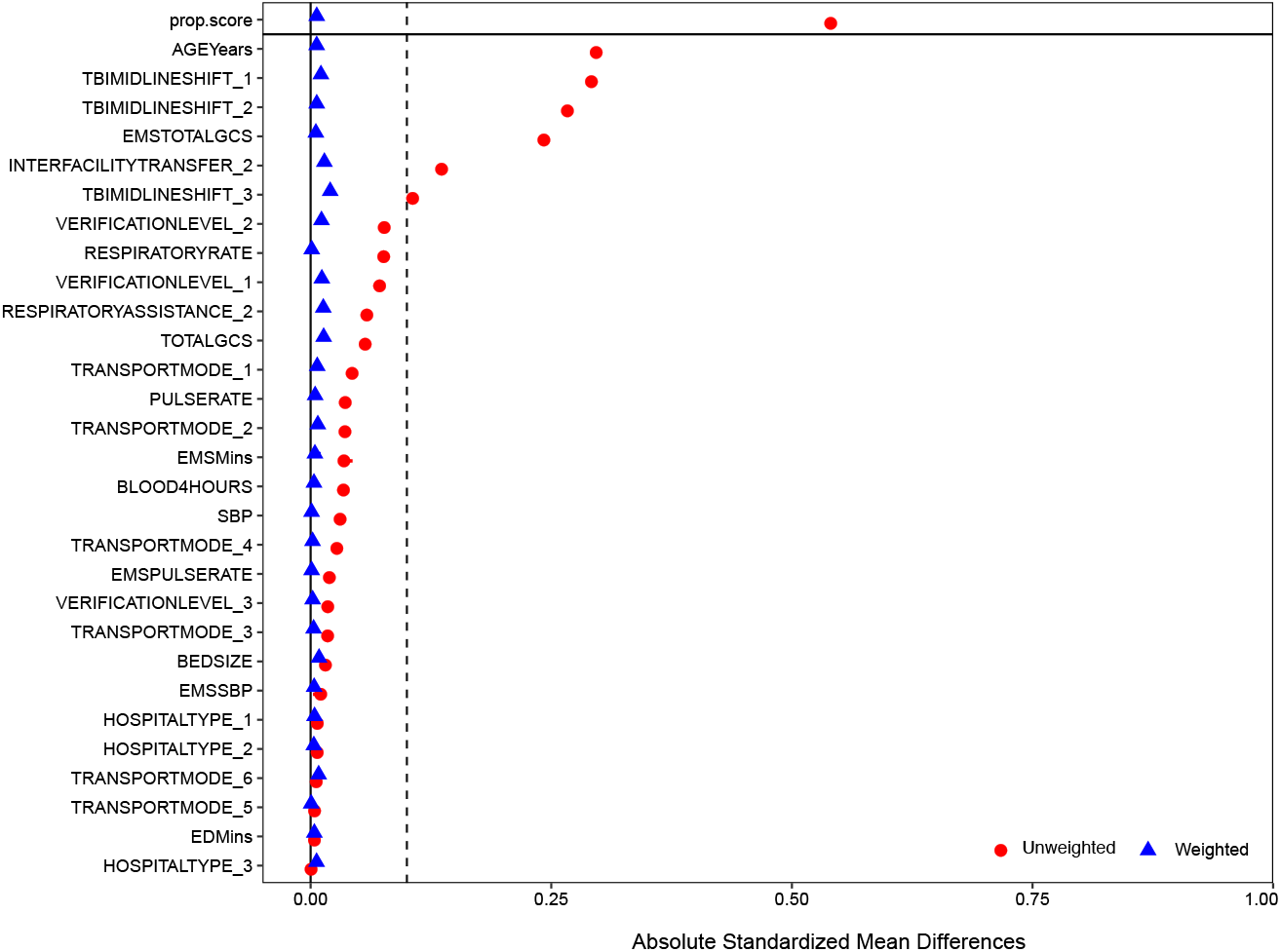
The ASD values of the covariates after adjustment: the dashed lines indicate the threshold of balancing.

### Estimation of ATE

The main research question in evaluating ICP monitoring in acute care TBI was: could ICP monitoring lower the mortality rate in TBI patients significantly in comparison with if no ICP monitoring was implemented? Before applying IPTW, the observed mortality rate in treated patients was 53.97% while the mortality rate in untreated patients was 56.34%. The ASDs results suggested the original unadjusted data were biased and potential significant confounding variables existed. Therefore, a direct comparison of mortality rates without adjustment was not valid. Furthermore, conventional logistic regression modeling would only suggest an association, not a causal relationship between ICP monitoring and lower mortality rates. By IPTW, the ATE of ICP monitoring in lowering the mortality rate after adjustment was -1.43% (95% CI: -2.36%, -0.51%; p-value=0.00238). Thus, using the trauma medical records collected in the largest NTDB, we successfully obtained a causal conclusion that the use of ICP monitoring was effective in lowering in-patient mortality for patients with severe TBI. Based on the -1.43% ATE identified here, a total of 668 (95% CI: 238, 1,101) deaths could be prevented if those 46,689 TBI patients in the control group had received ICP monitoring instead.

## Conclusion

Missingness is a critical issue in the use of real-world data for causal inference in health care evaluation. Handling missing data properly could avoid the waste of potential valuable information as well as obtain a more valid inference.

In this study, we compared different strategies to deal with missing data in causal analysis. MI models for each covariates did not have much influence in the ATE estimations. When there was a high proportion of missing data, MICE method performed much better than the complete data analysis, suggesting that proper data imputation is necessary. We also evaluated the effect of ICP monitoring on mortality rate in TBI patients using an observational NTDB dataset. MI strategy was performed to handle missing data so that more observations were incorporated in the analysis. After imputing the missing data, the IPTW method was applied to balance the baseline covariates and estimate the ATE. The visualization diagnosis and absolute differences indicated a perfect balance among all observed covariates after adjustment. Finally we successfully draw a causal conclusion that ICP monitoring was associated with a significantly reduced risk of death compared with treatment without an ICP monitoring among severe TBI patients. Our findings validated the management guidelines and provided evidence to help clinicians make decisions in their practice.

## Data Availability

All data produced in the present study are available upon reasonable request to the authors.

https://www.facs.org/quality-programs/trauma/tqp/center-programs/ntdb

## Data Availability

All data produced in the present study are available upon reasonable request to the authors.

https://www.facs.org/quality-programs/trauma/tqp/center-programs/ntdb

## Acknowledgement

This work was funded in part by the National Institute of General Medical Sciences (NIGMS) of NIH under award number R01GM141279 and by the Eunice Kennedy Shriver National Institute of Child Health and Human Development (NICHD) of NIH under award number R01HD107280.

